# Predicting attention deficits and functional recovery after glioma resection through functional executive networks: insights from dynamic properties

**DOI:** 10.1101/2024.05.03.24306578

**Authors:** Francesca Saviola, Luca Zigiotto, Jorge Jovicich, Silvio Sarubbo

**Affiliations:** CIMeC, Center for Mind/Brain Sciences, University of Trento, Rovereto (Trento), Italy; Department of Medical and Surgical Specialties, Radiological Sciences and Public Health, University of Brescia, Brescia, Italy; Department of Neuroscience, Division of Neurosurgery, S.Chiara Hospital, APSS Trento, Italy; Structural and Functional Connectivity Lab, S.Chiara Hospital, APSS Trento, Italy

**Keywords:** [gliomas, intrinsic dynamic functional connectivity, functional recovery, postsurgical attentional deficit]

## Abstract

**Background:** Postoperative short-term attentional and executive dysfunctions are common after brain tumor resection, significantly impacting patients’ quality of life and functional recovery. The current study investigated whether presurgical functional dynamics of key brain networks supporting executive functioning could predict postoperative neuropsychological outcomes.

**Methods:** Twenty-two patients with gliomas underwent longitudinal resting-state fMRI scans (before and three-months after surgery), along with neuropsychological assessments (before, one-week and three-months after surgery). Co-activation patterns analysis (CAPs) characterized the functional dynamic properties of executive networks, including the Fronto-parietal (FPN) and Dorsal Attention networks (DAN). Temporal network properties were examined for stability, integration, and centrality over timepoints. Partial least squares analyses and linear models explored associations between network dynamics and cognitive functioning.

**Results:** Immediate post-surgical attentional deficits were linked to pre-surgical FPN properties revealing associated dynamic patterns of network activation. Pre-surgical FPN temporal properties predicted not only immediate appearance or persistence of post-resection deficits, but also the longitudinal progression of attentional performance otherwise neglected. However, regardless of the severity of attentional deficit, at three months post-surgery, temporal properties and neuropsychological profiles did not significantly differ from the pre-surgical ones, indicating recovery to baseline beyond treatment strategies.

**Conclusions:** Our study demonstrates that presurgical dynamic properties of intrinsic executive networks alone can predict short-term postoperative neuropsychological outcomes, highlighting the clinical utility of temporal functional connectivity. These findings emphasize the potential for using intrinsic brain activity dynamics as predictive markers for postoperative recovery and planning tailored rehabilitation interventions for cognitive deficits.

**Key messages:**

- **What is already known on this topic:**
  - Attentive deficits are common in short-term post-glioma neurosurgery, yet their anticipation remains a challenge.
- **What this study adds:**
  - Integrating presurgical intrinsic fMRI dynamic connectivity properties within executive networks with pre-surgical attention scores enables the prediction of the severity of attentive deficits post-surgery.
- **How this study might affect research, practice or policy:**
  - The proposed prognostic framework has the potential to significantly impact patient outcome in neuro oncological treatments by informing the planning of tailored interventions, both at the level of pre-treatment and for post-treatment rehabilitation planning.

## Introduction

Postoperative attentional and executive dysfunctions are common following brain tumor resection, affecting both patients’ quality of life and functional recovery^1,2^. Indeed, after surgery immediate outcome in attentive domain in glioma patients can sometimes reveal cognitive deficits, even in absence of clear presurgical insufficient performance^3^. Importantly, also during awake brain surgery correct monitoring of several manifestations of attention and executive functions is challenging^4^. Therefore, the ability to predict the occurrence of this type of post-surgical deficits could be crucial for improving clinical practice, not only for surgical treatments, but also for further treatments more focused on illness progression such as radiotherapy^5^. Despite advancements in the field, accurately predicting temporary or permanent post-surgical deficits or recovery in attention and executive function at individual level remains demanding with current technology.

Large-scale functional brain networks are known to be involved in executive function processes, such as attention, working memory, inhibitory control, cognitive flexibility and shifting. Previous studies, both in healthy^6,7^ and glioma patients^8–11^ demonstrated how functional executive connections can sustain cognitive performance in the attentive domain. Specifically, two main neural networks seemed to be involved in several executive and attentive performances: fronto-parietal network (FPN) and dorsal attention network (DAN). The former, is considered a flexible network strictly related to goal-directed behavior or cognitive control^12^ and the latter is believed to be engaged during integrative externally directed attentional tasks^13^. Attentional functional networking can be reconstructed by looking at statistical dependencies in activity between distant but functionally interrelated regions in the brain, the so-called intrinsic functional connectivity^14^. Furthermore, given the inherent brain properties of dynamism, recent frameworks exhibited how spatiotemporal fluctuations of blood oxygenation level dependent (BOLD) signal strongly characterize time-varying properties of cognition at rest^15^, posing it as an extremely relevant methodology in the context of non-stationary cognitive shifting abilities such as attention. Consequently, longitudinal dynamic plastic changes of these intrinsic functional networks could be informative about the resilient support of cognitive performance, especially the attentive one.

In this study, we took advantage of dynamic intrinsic functional connectivity estimations, previously demonstrated to be clinically relevant^16^. We aimed at characterizing pathological functional temporal profiles of attentive networking and testing their potential as a clinical tool to predict these cognitive performance’s changes.

### Materials and Methods

#### Participants

A total of 22 gliomas patients were recruited at Santa Chiara Hospital in Trento, Italy. Approval for this study was obtained by the Ethical Committee of the Azienda Provinciale per i Servizi Sanitari (APSS, Neusurplan project, authorization ID A734). Subjects (16 males, age 47.8 ± 14.8 years, 11 right-hemisphere tumors) had a diagnosis of gliomas of different malignancy grades, with 13 high-grade (HGG) and 9 low-grade gliomas (LGG) patients^17^. See Table 1 for more demographic and clinical information and Supplementary materials for surgical procedure.

**Table 1:** Demographic and clinical characteristics of the sample.

| Patients | Brain tumor types |  |  | Statistic | p-value |
| --- | --- | --- | --- | --- | --- |
|  | Low grade gliomas | High grade gliomas | Total |  |  |
| Age (mean ± SD) | 46 ± 15 | 47 ± 15 | 48 ± 15 | t-value:-1.87 | 0.07 |
| Gender (# males) | 7 | 9 | 16 | $\chi^2$: 0.43 | 0.67 |
| Tumor lateralization (# Left) | 6 | 5 | 11 | $\chi^2$: -1.29 | 0.21 |
| Tumor WHO grade (# patients) | [grade-1: 1;<br>grade-2: 8] | [grade-3: 4;<br>grade-4: 9] | N.A. | N.A. | N.A. |
| Tumor volume cm<sup>3</sup> (mean ± SD) | 28.2 ± 29.9 | 32.6 ± 30.2 | 28.1 ± 28.9 | t-value=-1.74 | 0.09 |
| Awake surgery (# patients) | 9 | 8 | 17 | N.A. | N.A. |
| IDH-1 mutation (# patients) | 0 | 2 | 2 | N.A. | N.A. |
| MGMT methylation (# patients) | 0 | 6 | 6 | N.A. | N.A. |
| Radiotherapy [(# patients (week mean, SD))] | 0 | 9 | 13 | N.A. | N.A. |
| Proton therapy [(# patients (week mean, SD))] | 0 | 4 | 4 | N.A. | N.A. |
| Chemotherapy [(# patients (week mean, SD))] | 0 | 12 | 12 | N.A. | N.A. |

#### Dynamic network analysis

Resting-state fMRI (rs-fMRI) and structural T1-weighted images were acquired at Santa Chiara Hospital in Trento, Italy^18^. Following standard preprocessing steps (supplementary materials), we investigated longitudinal changes in intrinsic executive networking using a framewise dynamic functional connectivity method known as co-activation patterns (CAPs)^19^. This approach avoids assumptions about the hemodynamic response function for brain state estimation. Focused on executive networks, we estimated CAPs using predefined seed sets from literature^20^: Fronto-parietal network (FPN) and Dorsal attention network (DAN). Concatenating pre- and post-surgical timepoints, we extracted co-activation patterns in BOLD signals relative to the seeds, with temporal clustering detecting network fluctuations. Top 10% activation time points were selected, and consensus clustering determined k=4 as optimal for both seed sets. K-means clustering generated spatial z-maps for CAPs. Temporal metrics including IN-degree, OUT-degree, resilience, betweenness centrality, and occurrences were computed for each CAP across sessions^21,22^. Longitudinal changes (Δ) were assessed between post- (3-months later) and pre-surgery values.

#### Neuropsychological assessments

Each patient underwent a longitudinal neuropsychological assessment focusing on executive function domains at various time points^4,23^: presurgical, one-week post-surgery, and three-month follow-up. Attentional and executive function scores, obtained from attentional matrices and Trial Making Test (A, B, B-A scores), were analyzed^24^. Cognitive deficit percentages^3,4^ are reported in Table S1. Assessment occurred before surgery, one-week post-surgery, and three months later, adjusted for age and education. Pathological scores were dichotomously classified based on presence/absence of deficits one-week post-surgery. Δ scores were computed as differences between pre- and post-surgery performance, later linked with neuroimaging findings (Supplementary materials).

#### Statistical analysis

Statistical analyses were performed using Matlab 2019b and R software (v3.6.0) (https://cran.r-project.org/bin).

Neuropsychological scores and temporal properties of dynamic networks were separately tested for differences across subjects between sessions with a two-sample paired t-test. Moreover, aiming at detecting the most relevant functional networks in determining the following neuropsychological performance, Δ of neuropsychological scores in executive functions were tested for correlation using Pearson’s coefficient with Δ of temporal occurrences of each CAPs (as the most simple and informative metrics about temporal dynamics^25^).

To explore the relationship between executive functional networks (FPN and DAN) and deficit scores, we employed partial least squares correlations. Initially, we analyzed correlation matrices between behavioral (neuropsychological scores) and brain variables (CAP temporal features) across normal and impaired groups. Singular value decomposition of concatenated matrices yielded correlation latent components (LC) through permutations and bootstrap sampling. LC comprised behavioral and temporal feature weights, illustrating their contributions to brain-behavior correlations. Additionally, confirmatory analyses were conducted to assess network involvement in cognitive performance prediction (see Supplementary Materials).

Additionally, linear and linear mixed models (lm, lme4 functions of R-package) were utilized to predict postoperative attentive performance based on pre-surgical dynamic properties of executive networking. Various models were examined to comprehend and anticipate patient behavior (supplementary materials).

## Results

### Longitudinal executive and attentive profiles

Supplementary Table 1 shows the group average executive and attentive scores at the various treatment stages. The two-sample paired t-tests revealed that immediately post-surgery (i.e. 1 week after surgery) patients significantly decreased (p<0.05; t-value_TMTA_: -2.4, t-value_TMTB_: - 3.6, t-value_TMTB-A_: -3.1) executive and attentive performance scores, regardless of tumor grade and hemisphere, whereas attentional matrices showed no significant changes. The three-month follow-up time point showed no significant differences in executive and attentive performance (p>0.05) relative to the presurgical stage, regardless of tumor grade and features.

### Longitudinal temporal dynamics of executive networks

The CAPs characteristic of all subjects and retrieved for k=4 using the FPN seed are displayed in Figure 1A, representing several states of the executive network: (i) CAP1_FPN_ is characterized by large activation in frontal regions together with parietal bilateral regions which typically represents the static network, whereas de-activation areas are localized in somatosensory a visual regions; (ii) CAP2_FPN_ shows activations mainly in dorso-lateral frontal regions combined with a larger temporo-parietal involvement; (iii) CAP3_FPN_ depicts activations in areas belonging of the dorsal attention networking in regions similar to CAP2_FPN_ while showing de-activations in occipital regions; (iv) CAP4_FPN_ is nicely reminiscent of strong activations within fronto-parietal regions, the so-called “central-executive”. Boxplots exhibit the temporal features of each FPN state (Figure 1) over the time course, both pre- and 3-month post-surgically. No statistically significant differences were found when comparing the longitudinal properties of each state within the FPN across subjects (p-value > 0.05).

**Figure 1:**
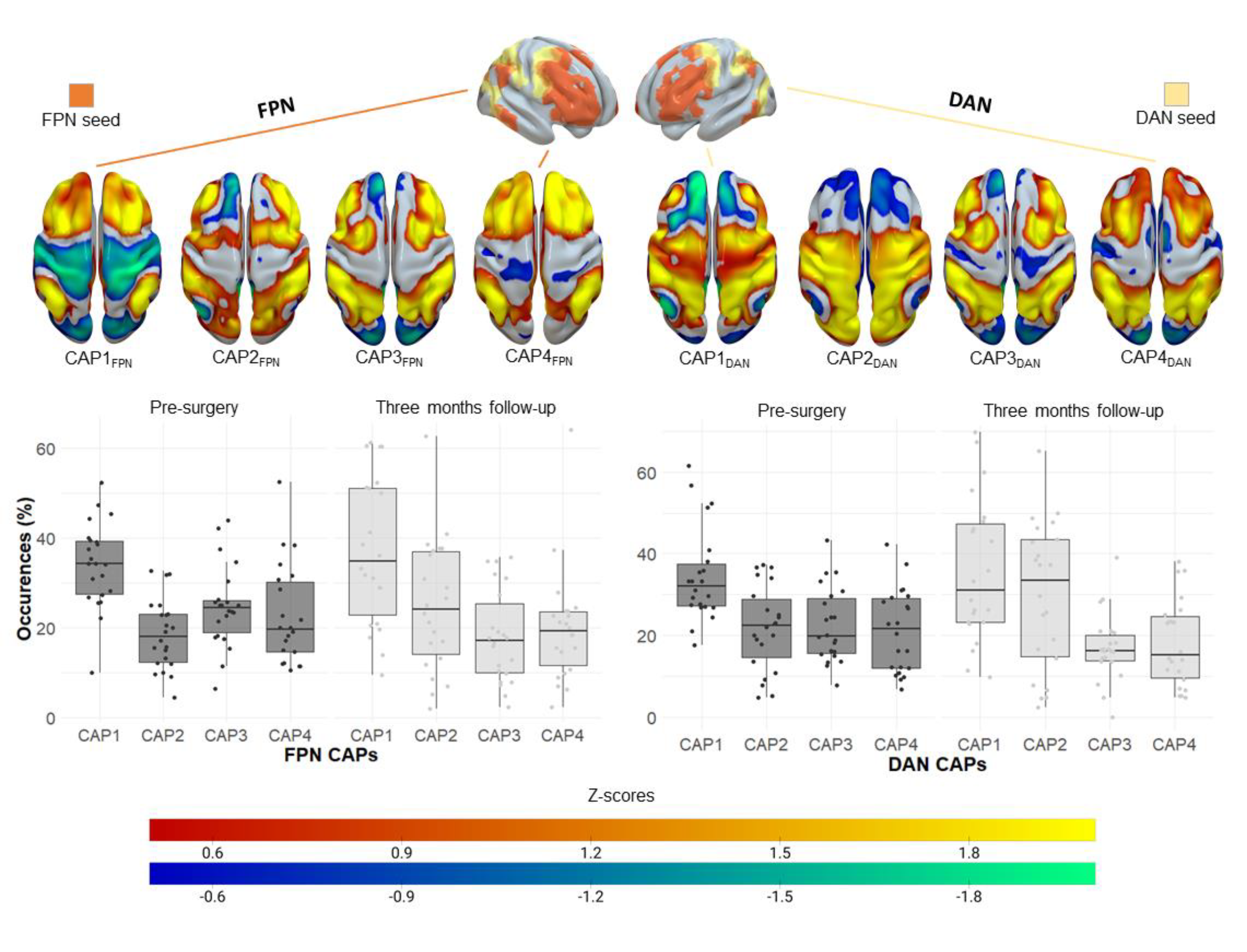
Longitudinal fronto-parietal and dorsal attention networks dynamism in gliomas. First row, anatomical placement of the fronto-parietal (FPN, left) and dorsal attention (DAN right) seeds from the seven networks parcellation (Yeo et al., 2011). Second row, four Co-activation patterns (CAPs) spatial distribution characteristics of all subjects for the two timepoints extracted from the corresponding seeds. Third row, boxplot of CAPs dynamic features over the time course (i.e. percentage of the time each subject is spending in each CAP over the time course).

The CAPs retrieved for k=4 using the DAN seed characteristic of all subjects are displayed in Figure 1B, representing several states of the executive network: (i) CAP1_DAN_ is characterized by large somato-sensorial activation within the temporal and parietal lobe with de-activation comprises within bilateral medial cingulate regions; (ii) CAP2_DAN_ is exhibiting a strong activation of temporo-parietal regions including also dorsal portion of the occipital lobe; (iii) CAP3_DAN_ is nicely reminiscent of the dorsal attention regions, with activation involving the intraparietal sulcus and the frontal eye-fields; (iv) CAP4_DAN_ depicts activations in similar regions of CAP3_DAN_ even if to a less extent in terms of strength. Boxplots exhibit the temporal properties of each DAN state (Figure 1) over the time course, both pre- and post-surgically. No statistically significant differences were found when comparing the longitudinal properties of each state within the DAN across subjects (p-value > 0.05).

### Temporal properties of fronto-parietal network are associated with attentional and executive cognitive profile

As previously shown, the pre-surgical and 3-month follow-up executive function assessments showed no significant differences among attentional deficit groups for each cognitive score (p>0.05). Only CAP4_FPN_, related to FPN, showed strict correlation with cognitive profile (Figure 2A). ΔTMT-B and ΔTMT-BA (3-month post-surgery vs. presurgical change) were negatively associated with CAP4_FPN_’s temporal duration. Negative ΔTMT scores indicated post-surgery cognitive deterioration, while positive or close to zero ΔTMT scores implied improvement or no change, respectively. Negative Δ of FPN temporal duration predicted impaired post-surgical cognitive performance, while positive or close to zero Δ indicated recovery at three-month post-surgery.

**Figure 2:**
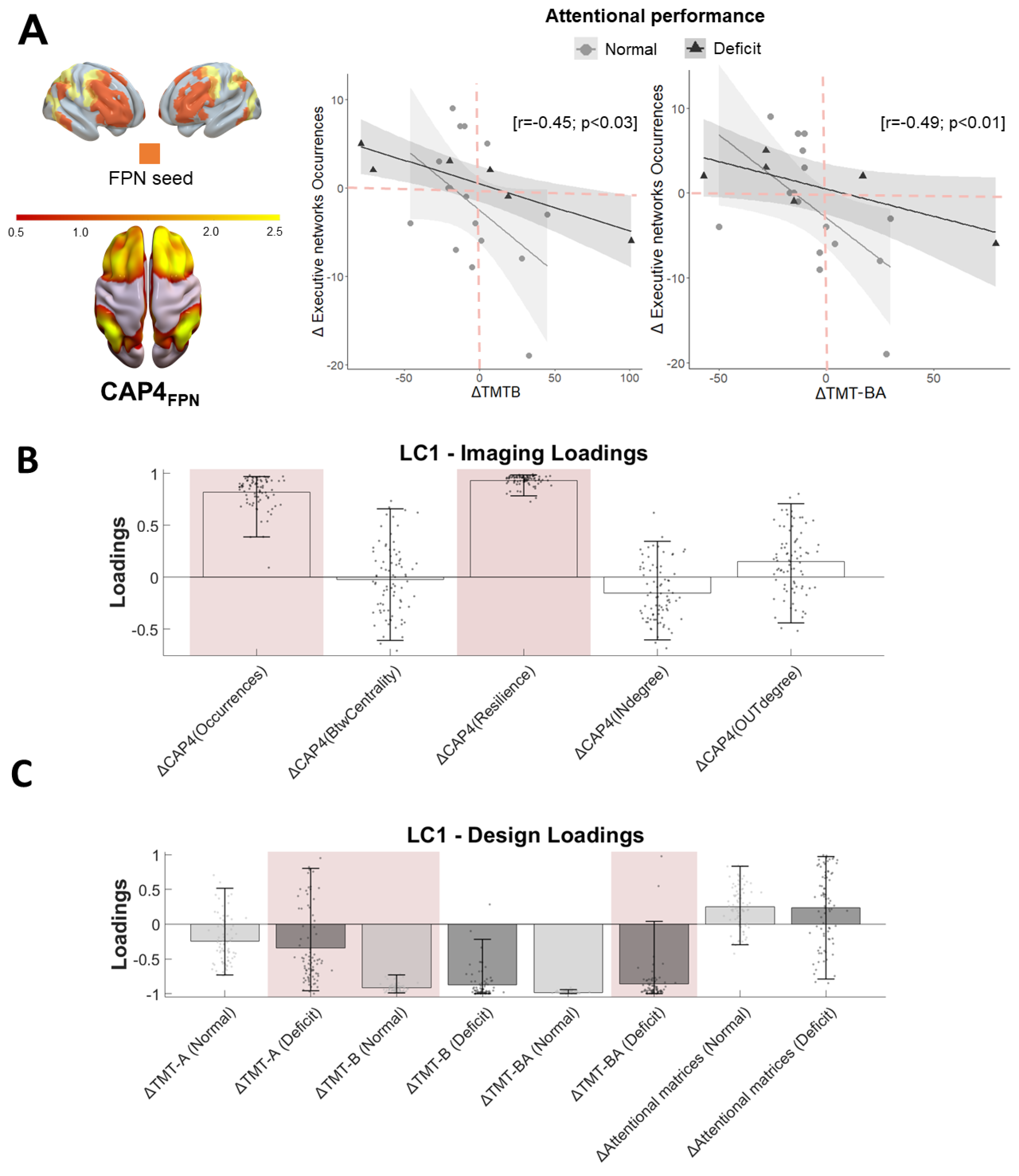
Functional signature of attentional cognitive outcome through fronto-parietal dynamism. A) Correlation between changes in attentional performance and changes in fronto-parietal CAP duration between the 3-months follow-up and the presurgical stages (ΔTMTB and ΔTMT-BA calculated by computing the difference between the performance scores between the evaluation after (3 months follow up) and before surgery). B) and C) Significant Latent Component (LC) identified with grouped behavioral Partial Least Squares Analysis explaining differing effects of covariance in patients with different attentive performance disregarding absolute group differences. Errorbars indicate bootstrapping 5th to 95th percentiles, robust results from permutation testing are highlighted by pink boxes.

To study how CAP4_FPN_’s temporal properties relate to overall attentional profiles, we employed behavioral grouped PLS. One significant latent component (LC; p=0.006; Table S2) highlighted a strong correlation (r=0.80) between behavioral and imaging scores (supplementary materials). CAP4_FPN_’s temporal feature weights, driving brain-behavior relationships across groups, are displayed in Figures 2B and 2C. Increased occurrences and resilience of CAP4_FPN_ were associated with lower ΔTMT-A in the attentive impaired group and with lower ΔTMT-B but higher Δ attentional matrices in the group with normal executive functioning. Further PLS analyses with FPN and DAN seeds revealed that CAP1FPN and CAP3_FPN_ also contributed to explaining ΔTMT scores (p=0.005; r=0.81), particularly for the FPN seed. For DAN, the significance of LC (p=0.014; r=0.78) in explaining behavioral patterns between groups was primarily attributed to higher Δ in CAP1_DAN_ in-degree, not specifically associated with a cognitive score (Supplementary materials, Figure S1, panel C and D).

### Prediction of attentional cognitive profile through temporal properties of fronto-parietal network

The initial linear mixed model assessed whether attentive performance could be predicted by longitudinal (3 months follow-up) dynamic properties of executive functional networks (Supplementary Materials, Model 1; Figure S2; Table S3). Attentive performance was defined by the neuropsychological scores previously described. No significant effects were observed for attentional matrices. However, all three TMT scores exhibited fixed significant effects (p<0.05): (i) presence of attentional deficit post-surgery in TMT-A and TMT-B; (ii) longitudinal betweenness centrality of dynamic FPN’s states in TMT-A and TMT-B. Significant interactions (p<0.05) were found between: (i) presence of attentional deficit and longitudinal dynamic properties of FPN’s state in all TMT scores; (ii) time, presence of attentional deficit, and longitudinal dynamic properties of FPN’s state in all TMT scores. These interactions underscored the importance of considering both the longitudinal aspect and the dual roles of executive network dynamism. FPN states’ stability properties were predictive of attention deficit presence with lower occurrences and higher resilience values. Additionally, the number of state transitions was relevant; higher IN- and OUT-degree values could explain attentional deficit presence.

Another linear model (Model 2, Table S4) was used to assess if attentional deficit presence could be predicted by presurgical executive network dynamic properties. Significant fixed effects (p<0.05) were observed for transition properties (IN- and OUT-degree). Subsequent specific models (Model 3, Table S5), examining individual neuropsychological scores, revealed additional network properties, particularly occurrences, relevant for prediction.

We investigated whether longitudinal attentive performance could be predicted by pre-surgical network dynamism. In the first model (Model 4, Figure 3; Table S6), we aimed to predict immediate post-surgical cognitive outcomes. Significant fixed effects (p<0.05) were found for IN-degree, with higher values linked to poorer performance. Similarly, IN-degree and OUT-degree FPN transitions were significant in the second model predicting attention performance at three months follow-up (Model 5, Figure 3; Table S7), particularly for executive scores (TMT-B and TMT-BA).

**Figure 3:**
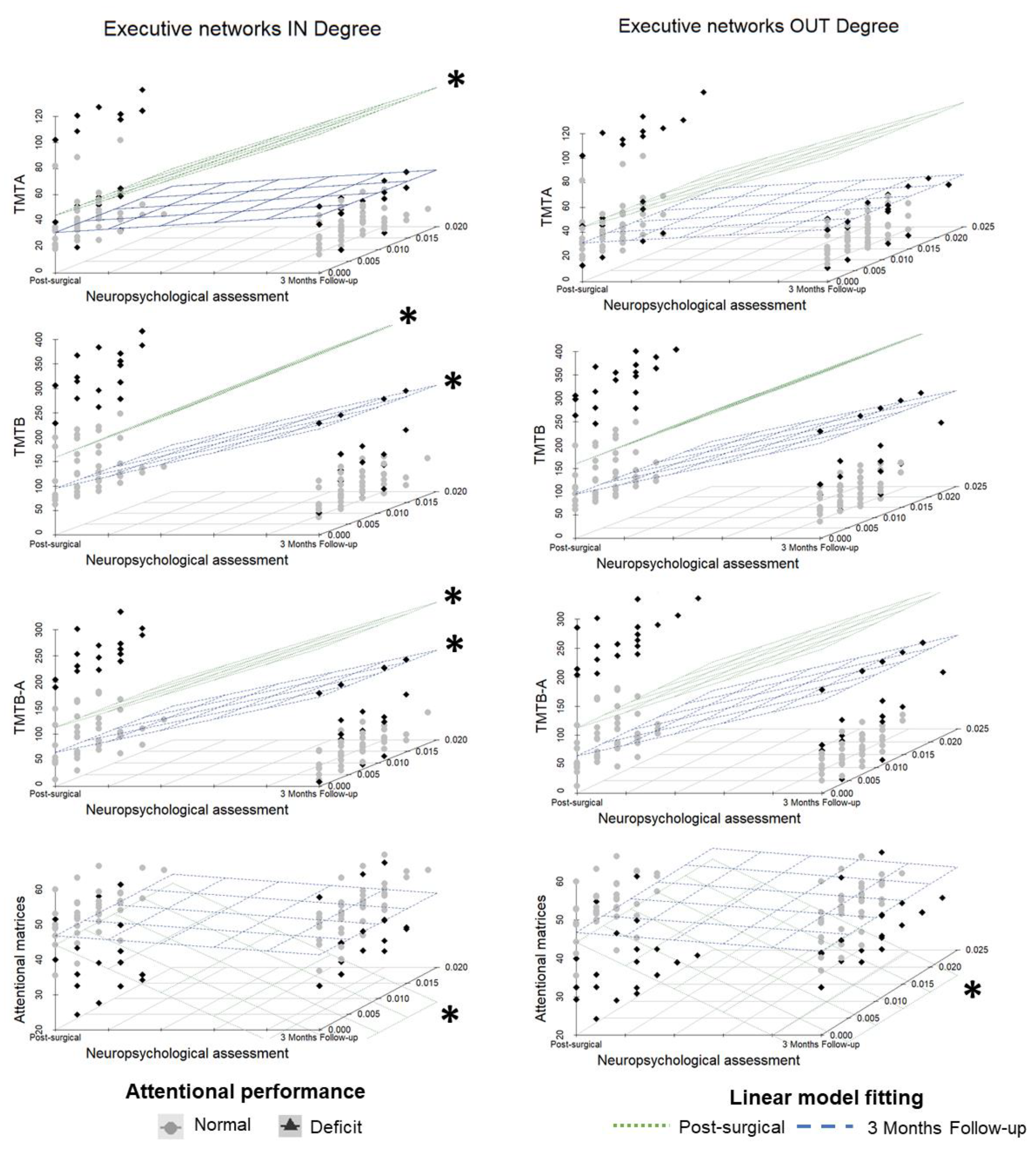
Prediction of postoperative (immediate and 3 months follow-up) attentional cognitive outcome through pre-surgical fronto-parietal temporal properties. 3D Scatterplots of linear models investigating respectively if presurgical dynamic properties of FPN could predict immediate post-surgical (model 4) or three-months follow-up (model 5) attentional and executive performance, displayed as a function of IN Degree properties of FPN (probability of transitions from other dynamics states to the FPN, first column) and OUT Degree properties of FPN (probability of transition from the FPN towards other dynamic states, second column) for each neuropsychological test. Significant model effect visualized as 3D plane are marked with *. The *Deficit* subgroup are patients with at least one deficit in the neuropsychological battery at 1-week post-surgery discharge, the *Normal* group has no deficits.

Finally, we explored whether pre-surgical behavioral and functional data alone could distinguish patients with or without attention deficits one-week post-surgery (Figure 4). Pre-surgical attentive and executive scores were normalized and examined alongside pre-surgical functional dynamic metrics (IN-degree and OUT-degree). A distinct distribution was observed between patients with post-surgery attention deficits and those without, suggesting a notable group difference. Short-term post-surgery reduction in attentive cognitive scores strongly correlated with pre-surgical increases in both IN-degree and OUT-degree indices of the FPN

**Figure 4:**
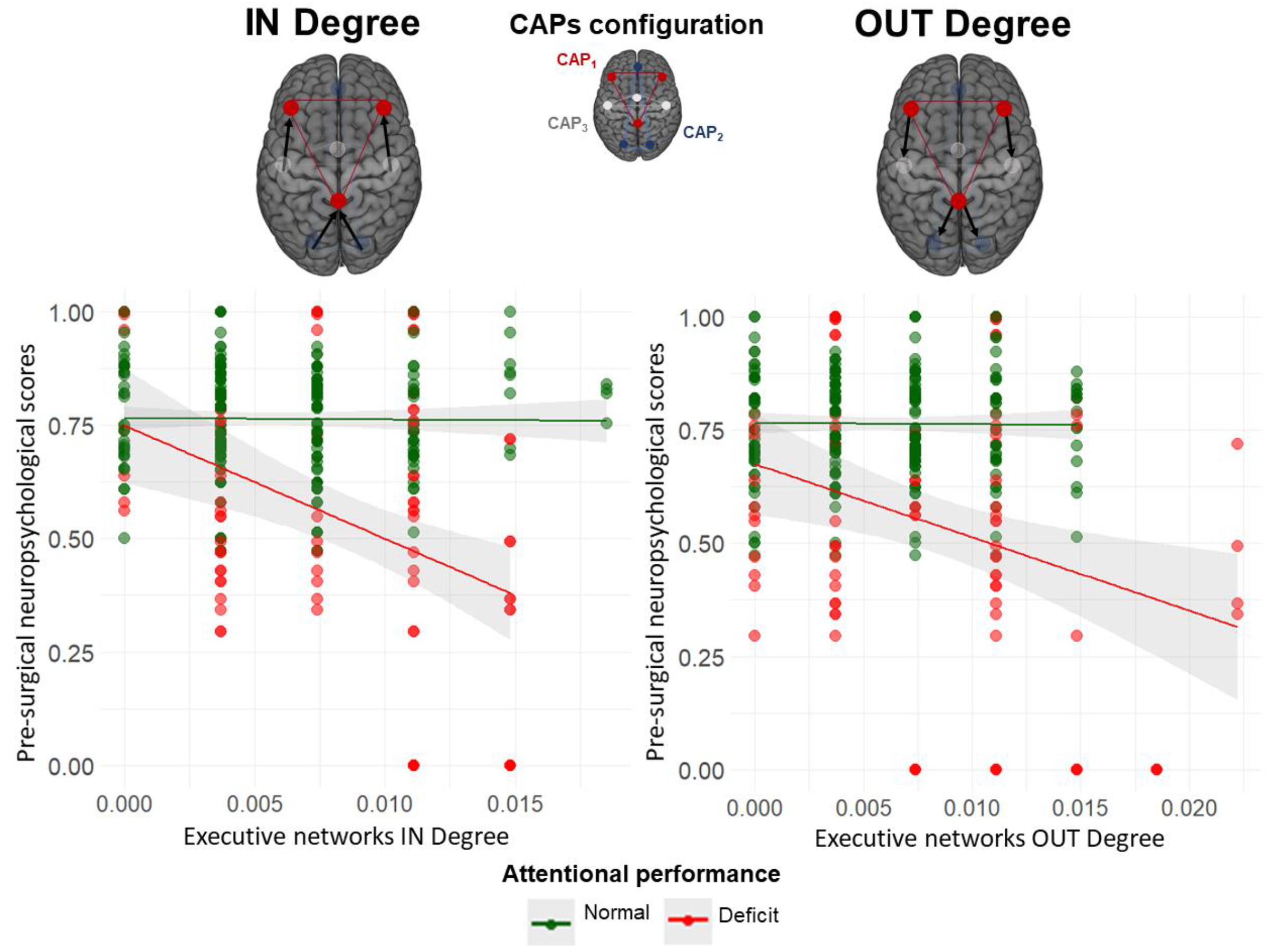
Pre-surgical data as indicator of post-surgical attentional deficits. The scatterplots depict the association between pre-surgical dynamic rs-fMRI properties of the FPN (x-axes) and pre-surgical performance in attentional and executive tasks (y-axis). FPN dynamics are represented by IN Degree properties (probability of transitions from other dynamic states to the FPN, left) and OUT Degree properties (probability of transitions from the FPN towards other dynamic states, right). Neuropsychological test scores are presented in normalized z-score forms. The colored dots distinguish two groups of patients: those who, 1-week post-surgery, showed deficits in at least one neuropsychological test (red dots) and those without deficits (green dots). Separate linear fits of pre-surgical data in these two groups are shown.

## Discussion

In this study, we investigated dynamic features of intrinsic executive and attentive network brain activity in glioma patients, focusing on the identification of presurgical functional signatures with a predictive value of post-surgical attentive deficits, extremely relevant for patients’ recovery and to drive surgical and non-surgical oncological treatments.

To the best of our knowledge, this is the first study investigating associations between large-scale executive and attentional brain network dynamics and post-surgery attentional deficits in gliomas. We took advantage of CAPs framework^22^ to estimate dynamic brain functional properties and move beyond static functional connectivity analysis of resting-state activity^26^. We revealed gliomas alterations of brain networking related to attentive deficits, especially associated with stability of the executive network, and employed them as predictors of post-surgical cognitive performance recovery.

### The glioma pathophysiology: implications in attention and executive dynamic brain networks

Previous studies showed how preserving the integrity of functional brain networks is crucial for recovery of several cognitive systems and abilities after surgery such as memory^26^. Individuals with gliomas have been largely studied in terms of static functional connectivity reorganization^27,28^ and plastic longitudinal changes after surgery^11,29,30^. However, only few studies have investigated the dynamic nature of plastic changes of resting-state networking in gliomas, and these studies have been cross-sectional in nature, comparing glioma patients to healthy controls^31^ or across different tumor grades^32^, reporting a global decrease in connectivity strength, occurrences and networks’ segregation. Yet the longitudinal evolution of functional dynamic networks, before and after glioma resection, remains largely unexplored, especially with regards to their predictive potentials for predicting treatment outcome.

In our study, we investigated within-subject longitudinal changes of functional dynamics of specific networks associated with attentional performance such as FPN and DAN. We found that, besides the presence of immediate post-surgical attentional deficits (which recover for the vast majority of the patients at 3-month follow-up), single-subject network dynamics do not significantly change between presurgical and 3-month follow-up states. This may subtend the idea that overall longitudinal dynamic properties of FPN and DAN are not affected per se by the surgical resection, as previously reported by static connectivity analyses^26^. However, the investigation of associations between longitudinal temporal properties variation and changes in cognitive functions revealed that a strongly co-activated FPN state (CAP4_FPN_) was correlated to longitudinal executive control performance, implying an involvement of this network dynamics in sustenance of cognitive performance^33^.

In this context, investigating also the appearance of relative longitudinal changes in attentive performance, moving beyond the simple recovery of pre-surgical impairment, might be extremely beneficial on several perspectives. First, from the immediate post-surgical time point, patients who persist or who start experiencing attentional deficits at the current-state of the art are typically not treated with rehabilitation, given the natural recovery of the impairment. Our findings, by predicting at the presurgical time point either the onset or the maintenance of post-surgical attention deficit, would allow the planning of a selected and selective cognitive treatment from the very beginning of patients’ clinical care. The well-time and effective treatment proposed for patients who might experience attentive deficit would potentially allow a faster functional cognitive recovery enabling a sooner return to work for patients and an improved quality of life. Moreover, this would reflexively impact not only clinical cost, by providing a specific focus on patients needing treatment, but also a less intense follow-up for patients with a predicted positive cognitive outcome. Furthermore, our data clearly report worsening in cognitive performance one week and 3 months after surgery for several patients (Fig. 2A). This result, even in absence of pathological performance, represents an impairment in patients’ perception of Health-Related Quality of Life (HRQoL). Therefore, attentive worsening prediction, as well as early interventions, could significantly improve patients’ HRQoL.

### Stability of the intrinsic fronto-parietal network in glioma: a potential marker of post-surgery compensation

The presence of immediate post-surgical attentional deficits, as evidenced also in executive functioning (i.e. TMT-A, TMT-B and TMT-BA), were negatively associated with changes in both occurrence and resilience properties of one specific FPN state relative to presurgical stage. A reduction of FPN occurrences has been previously reported both statically and in other types of dynamic connectivity analysis in gliomas while comparing hemispheric plasticity or relative to healthy populations^31,34^. Furthermore, studies examining dynamic connectivity and cognitive performance in glioma patients have highlighted that the absence of stable reconfiguration properties within FPN network serves as a negative prognostic indicator for functional recovery^35,36^. Taken together, these results suggest that plastic reorganization of FPN temporal properties, in terms of longitudinal changes of network occurrences, might be strictly associated to changes in patient’s cognitive performances. Indeed, greater longitudinal changes of FPN networking stability (i.e. higher Δ of occurrences or resilience) in patients with attentional deficits may describe worsening cognitive trends where patients do not change their cognitive performance over time, impairing recovery. This will reinforce the idea that an aberrant dynamic reconfiguration of FPN, especially its temporal stability^37^, may result in impaired cognitive compensation 3-month post-glioma resection. The FPN is considered as a key hub for cognitive flexibility, task-shifting and goal-oriented behavior; therefore, our findings support the idea that modifications in its temporal properties might impact cognitive executive functioning and plasticity.

### Functional dynamics of pre-surgical intrinsic executive networks predicts post-surgical attentive deficits

However, besides the known association between FPN activity and cognitive executive performance, prognostic attentive outcomes after surgery have been scarcely investigated. In this study, we also demonstrated how favorable attentive performance can be predicted from cognitive dynamism of executive networks derived from presurgical resting-state fMRI. Recently, negative evidence in favor of an anatomo-functional pre-surgical predictor of executive dysfunction was reported^38^. Therefore, aiming at a better understanding of the topography of executive function performance we looked at longitudinal plastic reorganization of temporal network properties of FPN. The richness of network dynamic approach enables us to characterize pre-surgical patterns contributing to the short-term post-surgical attentional deficit in gliomas that goes beyond its pure biological features represented by clinical characteristics. Indeed, we demonstrated that longitudinally attentive and executive functions (i.e. especially TMT component) can be predicted by the interaction between time, attentional deficit presence and temporal FPN properties. Furthermore, presurgical intrinsic network dynamic properties of FPN were predictive of post-surgery attention deficits or performance, both immediately after surgery and at 3-month follow-up. In this context, we emphasize the significance of two key aspects of FPN dynamics: stability (as indicated by occurrences) and temporal flexibility (represented by IN- and OUT-degree) in determining pre-existing attentional deficits and their persistence post-surgery. Specifically, a less stable and constantly switching FPN network is associated with attentional deficits that are present before surgery and continue to persist afterward. Importantly, our results are unrelated to tumor grade (i.e. high- or low-grade) neither to lateralization (i.e. left or right brain hemisphere): this crucial result reinforce the idea and the need to definitively leave behind the concept of localizationism in order to consider brain and its deficit as a connectome, with the crucial role of cortical networks and subcortical connections rather than lesion or tumor location^39^.

Our results provide novel and clinically relevant insights that can inform treatment options for patients. Considering the reported relevance of FPN networking, it is paramount to factor it into decisions regarding therapeutic options to minimize patient’s post-surgery recovery time. Indeed, predicting and recognizing that patients with specific pre-surgical FPN dynamic characteristics should not experience attentional deficits or should quickly recover can provide valuable guidance for radiotherapists. This information can help them determine the appropriate radiation dose to administer near the surgical cavity and tumor.

On the other hand, a more targeted radiant treatment, focusing specifically on the surgical cavity while minimizing exposure to neighbouring regions, should be thoroughly evaluated for patients at risk of retaining post-surgery deficits. Additionally, our study provides valuable predictive information about a patient’s cognitive status following surgery, which can be crucial for their understanding and preparation.

Further longitudinal studies are essential to validate the subject-specific predictive potential of post-treatment cognitive outcomes derived from pre-treatment intrinsic dynamic functional connectivity properties.

## Conclusions

Taken together, in this work we demonstrate that post-surgical attentive performance of patients who underwent glioma surgical resection is related to presurgical intrinsic network dynamic properties, in particular the executive network, such as FPN, irrespective of glioma grade or lateralization. The observed deficits in executive functions may be attributed to alterations in temporal persistence of highly co-activated frontoparietal network nodes. These findings suggest that interventions aimed at restoring the robust occurrence of this functional network, potentially through techniques such as brain stimulation, could pave the way for preventing or recovering attentional performance impairments resulting from surgery in glioma patients as well as following radiant treatment, which can, at least temporarily, impair attention and memory functions^40^. Furthermore, these results strengthen the importance of investigating pre-surgical intrinsic dynamics of complex networks supporting higher cognition to improve patient’s prognosis and optimize patient outcomes.

## Funding

Autonomous Province of Trento, Italy: Project: “NeuSurPlan and integrated approach to neurosurgery planning based on multimodal data” to S.S. and J.J.

Italian Ministry of Education, University and Research: Dipartimento di Eccellenza project 2018-2022 to F.S.

Lucarelli Irion Foundation, Rovereto, Italy, who supported F.S.

## Conflict of Interest

None declared.

Authorship

## Francesca Saviola

Writing - original draft, Formal analysis, Visualization, Software, Investigation, Writing - review & editing. **Luca Zigiotto:** Writing - original draft, Formal analysis, Project administration, Writing - review & editing. **Jorge Jovicich:** Supervision, Writing - review & editing, Investigation. **Silvio Sarubbo:** Funding acquisition, Project administration, Writing - review & editing, Supervision, Conceptualization.

## Data availability

Data will be made available in de-identified format upon request to the authors.

## Acknowledgments

We would like to thank Ludovico Coletta and Manuela Moretto for their insightful comments and suggestions about the methodology.

