## Supplementary Materials for "Predicting attention deficits and functional recovery after glioma resection through functional executive networks: insights from dynamic properties"

### Supplementary materials and methods

#### Surgical procedure

Volumetric T1 with gadolinium and T2/Flair (for LGG) images, merged with the tractography and functional reconstructions of the critical cortico-subcortical structures, was used for neuro-navigation for each patient both in awake surgery and in general anaesthesia. For awake surgery, the cortical and subcortical mapping was performed at 60Hz, 1ms duration and 2-4mA of amplitude, as previously reported[1,2]. The threshold was set eliciting speech arrest at the level of the ventral premotor cortex (VPMC), regardless of tumor laterality. During awake surgery a customized intraoperative neuropsychological monitoring was performed for each patient. Surgical resection was therefore stopped when functional responses were elicited from the cortical and subcortical stimulation of eloquent structures. Depending on the lesion lateralization and location, the following tasks were performed during awake surgery: counting (0-10) and motor task, object naming, verb generation, reading and comprehension, palm-pyramid-tree test (PPTT), Stroop test, line bisection, the modified version of “reading the mind in the eyes", as previously reported[1–5].

#### MRI acquisition

Rs-fMRI scans were acquired with a 2D T2*-weighted gradient-echo echo-planar imaging (EPI) sequence (TR = 2600 ms, voxel resolution = 4×4×4mm3, TE = 45 ms, FA = 87°, FOV = 256 × 256 mm2, number of slices = 33-35, acceleration factor ASSET = 2, AC-PC acquisition) lasting approximately 12 minutes (275 volumes). Participants were instructed to keep their eyes open and stay still for the entire duration of scanning session. Anatomical images were acquired with a structural T1-weighted scan (IRGE, TR = 10.6 ms, voxel resolution = 0.5 × 0.5 × 1.0 mm3, TE = 4.23 ms, TI = 450 ms, FA = 12°, FOV = 256 × 256 mm2, number of slices = 156-192, acceleration factor ASSET = 2).

#### MRI pre-processing

The preprocessing steps of rs-fMRI were preceded and followed by off-line quality check through visual inspection of temporal signal-to-noise ratio and standard deviation maps (for a detailed explanation of the steps see previous work on the same population[6]). Rs-fMRI and structural T1-weighted images were preprocessed with the Statistical Parametric Mapping SPM12 software (https://www.fil.ion.ucl.ac.uk/spm/software/spm12/). After DICOM to Nifti conversion of all scans, the first 10 seconds of the rs-fMRI acquisition were removed for a steady state signal. Pre-processing included the following steps: (1) slice timing and head motion correction; (2) co-registration of the T1-weighted anatomical image to the rs-fMRI time series; (3) T1-weighted image segmentation to grey matter, white matter and cerebro-spinal fluid; (4) rs-fMRI temporal filtering (median, 4th order linear detrending and 2nd order low pass filtering, Butterworth f<0.1 Hz); (5) regression from the rs-fMRI time series of 6 head motion parameters and white matter and cerebro-spinal fluid signals; (7) normalization to standard MNI template space; (8) spatial smoothing with 2 voxels Full Width Half Maximum Gaussian kernel size.

#### Neuropsychological assessment

The percentage of cognitive deficit, i.e., pathological scores, are reported for every test in Table S1. Neuropsychological assessment was conducted with a validated battery of tests widely used for LGG and HGG[1,7,8] and performed before surgery (12.96±9.34 days), 1 week after surgery (before hospital discharge), subsequently after 3 months before or after MRI scanning to be associated with neuroimaging measures. Neuropsychological scores were adjusted for age and education (following[8]); pathological scores, to be later associated with dynamism of functional networks, were then calculated using a dichotomous classification based on the presence or absence of a cognitive deficit at the evaluation before hospital discharge (i.e. 1 week after surgery) in at least one of the neuropsychological scores, i.e. a score under cut-off (pathological) or in the normal range of performance.

#### Partial Least Squares Analysis

To better understand the relationships between the executive FPN and DAN CAP's temporal measures and deficit scores in the executive domain, we applied two partial least squares correlations (PLSC, behavioral grouped version[9,10]).

We first computed an analysis with a correlation matrix between normalized "behavioral variables" (neuropsychological scores of executive functions) and normalized brain variables (only with CAP temporal features of the FPN states correlated with neuropsychological score) across groups defined based on the deficit in executive functioning [normal group and impaired group]. Correlational matrices were concatenated across subjects of different runs and singular value decomposition of this matrix led to the estimation of correlation latent components (LC) via 1000 permutations and 500 bootstrap samples. Each LC is further composed by a set of behavioral weights and FPN CAP temporal features weights, which represent how largely each variable contributes to the multivariate brain-behavior correlations across runs[11]. Moreover, two additional confirmatory PLS analyses were performed respectively between the FPN or the DAN four CAP's temporal measures and executive score across different groups to understand the overall involvement of networks in predicting cognitive performance

#### Linear models

Linear models were used to investigate the prediction of attentional and executive profile from the time-varying connectivity of the Fronto-parietal networking (FPN). In what follows each model is described by introducing each response and relative predictors.

1) Model 1: Linear mixed model
   The first model was used to test the hypothesis that dynamic longitudinal properties of FPN could predict the longitudinal attentive and executive performance of the patients. Therefore, for each attentional score (attentional matrices, TMT-A, TMT-B and TMT-BA) the following model was applied, using pre-surgical (as T0) and three months follow-up data (as T1). The presence of postsurgical attentional deficit was defined as a categorical variable described in the main text.

*Attentional scores ~ Time × Dynamic network properties × Attentional deficit postsurgical presence + (1|Subject)*

   Results of the model are displayed in Table S3.

2) Model 2: Linear model
   The second model was used to test the hypothesis that presurgical dynamic properties of FPN could predict the presence of attentional deficit in the immediate post-surgical timepoint. Therefore, by considering the presence of postsurgical attentional deficit (defined as a categorical variable described in the main text) as response, the following model was applied, using pre-surgical data only.

*Attentional deficit postsurgical presence ~ Presurgical dynamic network properties*

Results of the model are displayed in Table S4.

3) Model 3: Linear model
The third model was used to test the hypothesis that presurgical dynamic properties of FPN could predict attentional and executive pre-surgical performance. Therefore, for each attentional score (attentional matrices, TMT-A, TMT-B and TMT-BA) the following model was applied, using pre-surgical data only.

*Presurgical attentional scores ~ Presurgical dynamic network properties*

Results of the model are displayed in Table S5.

4) Model 4: Linear model
The fourth model was used to test the hypothesis that presurgical dynamic properties of FPN could predict immediate post-surgical attentional and executive performance. Therefore, for each attentional score (attentional matrices, TMT-A, TMT-B and TMT-BA) the following model was applied, using pre-surgical data only for the MRI data and immediate post-surgical data for the neuropsychological part.

*Postsurgical attentional scores ~ Presurgical dynamic network properties*

Results of the model are displayed in Table S6.

5) Model 5: Linear model
The fifth model was used to test the hypothesis that presurgical dynamic properties of FPN could predict three months post-surgical attentional and executive performance. Therefore, for each attentional score (attentional matrices, TMT-A, TMT-B and TMT-BA) the following model was applied, using pre-surgical data only for the MRI data and three months post-surgical follow up data for the neuropsychological part.

*Follow-up attentional scores ~ Presurgical dynamic network properties*

Results of the model are displayed in Table S7.

#### Supplementary Tables

Table S1: Age and education adjusted attention and executive cognitive scores of glioma patients at different time points of their intervention (Pre: pre-surgical, Post: immediate post-surgical, FU 3 m: follow-up at 3 months). The last column shows the % of patients with cognitive pathological scores in each test (i.e., relative to the normal population).

| Cognitive Tests | Time | Mean cognitive score (SD) | % of patients with deficit |
|---|---|---|---|
| Attentional matrices (selective attention) | Pre | 76.3 (±13.1) | 4.5 % |
| | Post | 73.7 (±17.5) | 13.64 % |
| | FU 3 m | 77.9 (±12.9) | 0.0 % |
| Trail Making Test part A (divided attention) | Pre | 30.8 (±19.7) | 4.5 % |
| | Post | 44.0 (±28.8) | 13.6 % |
| | FU 3 m | 31.1 (±12.0) | 0.0 % |
| Trail Making Test part B (alternating attention, mental/cognitive flexibility) | Pre | 100.4 (±59.7) | 4.5 % |
| | Post | 159.0 (±96.6) | 18.2 % |
| | FU 3 m | 94.9 (±51.9) | 0.0 % |
| Trail Making Test part B - A (alternating attention, mental/cognitive flexibility) | Pre | 69.5 (±42.6) | 4.5 % |
| | Post | 112.9 (±79.8) | 27.3 % |
| | FU 3 m | 64.3 (±44.2) | 0.0 % |

Table S2: **Bootstrap data corresponding to Figure 2**: PLSC results from attentional cognitive outcome through fronto-parietal dynamism. The table shows bootstrap mean and 5 to 95 percentiles of behavior weights and brain weights.

| Weights type | Item | Mean | 5th percentile | 95th percentile |
|---|---|---|---|---|
| **Behaviour:** | ∆TMTA (Normal) | -0.28 | -0.73 | 0.31 |
| **Attentive performence** | ∆TMTA (Deficit) | -0.90 | -0.99 | -0.58 |
| | ∆TMTB (Normal) | -0.98 | -0.99 | -0.95 |
| | ∆TMTB (Deficit) | 0.18 | -0.52 | 0.74 |
| | ∆TMTB-A (Normal) | -0.38 | -0.98 | 0.69 |
| | ∆TMTB-A (Deficit) | -0.87 | -0.99 | 0.29 |
| | ∆Attentional matrices (Normal) | -0.86 | -0.99 | 0.13 |
| | ∆Attentional matrices (Deficit) | 0.30 | -0.88 | 0.93 |
| **$CAP4_{FPN}$ properties** | ∆Occurrences | 0.85 | 0.53 | 0.98 |
| | ∆Betweeneess centrality | 0.04 | -0.52 | 0.62 |
| | ∆Resilience | 0.95 | 0.86 | 0.98 |
| | ∆In-degree | -0.12 | -0.62 | 0.39 |
| | ∆Out-degree | 0.19 | -0.28 | 0.72 |

Table S3.1: **Model 1**, Executive and attentive neuropsychological profile showing significant effects revealed by a linear mixed model of longitudinal changes (pre-surgical and three-months follow-up) in brain tumor patients stratified into two groups (with or without attentional deficit in the immediate post-surgical time point). The model included the following main predictors: *time* (a positive effect means neuropsychological score increases regardless the presence of attentional deficit and dynamic properties), *attentional deficit* (a positive effect means neuropsychological score increases regardless the time and dynamic properties), *time × attentional deficit* (a positive effect means that longitudinal neuropsychological score increases faster in patients with attentional deficit relative to normal one). The model also included dynamic functional connectivity scores of the Fronto-Parietal network reported in other tables. Significant ($p < 0.05$) fixed effects are emphasized in bold and an asterisk.

| ***Response*** | ***Predictors*** *[Estimate β, p-value]* | | | ***Random effects*** |
|---|---|---|---|---|
| | *Time* | *Attentional Deficit* | *Time × Attentional Deficit* | $\sigma^2$ |
| ***Attentional matrices*** | [-0.3, 0.9] | [-8.1, 0.1] | [2.4, 0.6] | 17.9 |
| ***TMT-A*** | [-0.6, 0.9] | [10.2, 0.2] | [-9.2, 0.2] | 55.9 |
| ***TMT-B*** | [80.8, 0.7] | [71.3, **0.0004]***] | [-32.7, **0.04**] | 272.4 |
| ***TMT-BA*** | [52.5,0.7] | [60.8, **0.001**]* | [-23.3, 0.1] | 159.4 |

Table S3.2: **Model 1**, Executive and attentive neuropsychological profile showing significant effects revealed by a linear mixed model of longitudinal changes (pre-surgical and three-months follow-up) in brain tumor patients stratified into two groups (with or without attentional deficit in the immediate post-surgical time point). The model included the following main predictors of dynamic functional connectivity patterns of the Fronto-Parietal Network: *betweenness centrality; occurrences, in degree, out degree and resilience.* For all of them, a positive effect means neuropsychological score increases regardless of time, other dynamic properties and the presence of attentional deficit. The model also included time and attentional deficit presence reported in other tables. Significant ($p < 0.05$) fixed effects are emphasized in bold and an asterisk.

| ***Response*** | ***Predictors*** *[Estimate β, p-value]* | | | | | ***Random effects*** |
|---|---|---|---|---|---|---|
| | *Betweenness centrality* | *Occurrences* | *In Degree* | *Out Degree* | *Resilience* | $\sigma^2$ |
| ***Attentional matrices*** | [0.5, 0.4] | [0.1, 0.4] | [-259.6, 0.1] | [-192.7, 0.2] | [-73.6, 0.6] | 17.9 |
| ***TMT-A*** | [-1.5, 0.2] | [-0.2, 0.1] | [229.9, 0.4] | [219.5, 0.4] | [415.9, 0.1] | 55.9 |
| ***TMT-B*** | [-4.9, **0.035]*** | [-0.2, 0.6] | [605.2, 0.4] | [600.9, 0.3] | [362.4, 0.5] | 272.4 |
| ***TMT-BA*** | [-3.9, **0.032]*** | [0.1, 0.7] | [292.6, 0.6] | [340.5, 0.4] | [-110.6, 0.8] | 159.4 |

<u>Table S3.3:</u> **Model 1**, Executive and attentive neuropsychological profile showing significant effects revealed by a linear mixed model of longitudinal changes (pre-surgical and three-months follow-up) in brain tumor patients stratified into two groups (with or without attentional deficit in the immediate post-surgical time point). The model included the following interaction predictors of longitudinal dynamic functional connectivity patterns of the Fronto-Parietal Network: *betweenness centrality* × *time, occurrences* × *time, in degree* × *time, out degree* × *time and resilience* × *time.* For all of them, a positive effect means that longitudinal neuropsychological score increases faster in time regardless of the presence of the attentional deficit. The model also included time and attentional deficit presence predictors reported in other tables. Significant ($p < 0.05$) fixed effects are emphasized in bold and an asterisk.

| ***Response*** | ***Predictors*** *[Estimate* β*, p-value]* | | | | | ***Random effects*** |
|---|---|---|---|---|---|---|
| | *Time × Betweenness centrality* | *Time × Occurrences* | *Time × In Degree* | *Time × Out Degree* | *Time × Resilience* | $\sigma^2$ |
| ***Attentional matrices*** | [0.6, 0.4] | [-0.1, 0.5] | [380.9, 0.1] | [253.9, 0.2] | [32.4, 0.8] | 17.9 |
| ***TMT-A*** | [0.9, 0.6] | [0.2, 0.5] | [13.7, 0.9] | [17.8, 0.9] | [-324.7, 0.3] | 55.9 |
| ***TMT-B*** | [2.0, 0.6] | [0.5, 0.4] | [-1047.3, 0.2] | [-937.0, 0.2] | [-424.9, 0.5] | 272.4 |
| ***TMT-BA*** | [1.1, 0.7] | [0.2, 0.7] | [-817.0, **0.02**]* | [-782.9, 0.2] | [2568.9, **0.002**]* | 159.4 |

Table S3.4: **Model 1**, Executive and attentive neuropsychological profile showing significant effects revealed by a linear mixed model of longitudinal changes (pre-surgical and three-months follow-up) in brain tumor patients stratified into two groups (with or without attentional deficit in the immediate post-surgical time point). The model included the following interaction predictors of longitudinal dynamic functional connectivity patterns of the Fronto-Parietal Network: *betweenness centrality* × *attentional deficit, occurrences* × *attentional deficit, in degree* × *attentional deficit, out degree* × *attentional deficit and resilience* × *attentional deficit.* For all of them, a positive effect means that longitudinal neuropsychological score increases faster in patients with attentional deficit relative to normal ones regardless of time. The model also included time and attentional deficit presence predictors reported in other tables. Significant ($p < 0.05$) fixed effects are emphasized in bold and an asterisk.

| ***Response*** | ***Predictors*** *[Estimate β, p-value]* | | | | | ***Random effects*** |
|---|---|---|---|---|---|---|
| | *Attentional Deficit × Betweenness centrality* | *Attentional Deficit × Occurrences* | *Attentional Deficit × In Degree* | *Attentional Deficit × Out Degree* | *Attentional Deficit × Resilience* | $\sigma^2$ |
| ***Attentional matrices*** | [0.6, 0.6] | [0.4, 0.1] | [-299.6, 0.3] | [253.9, 0.2] | [-365.8, 0.2] | 17.9 |
| ***TMT-A*** | [-5.0, **0.027]*** | [-1.3, **0.001]*** | [1726.2, **0.001]*** | [1819.4, **0.001]*** | [955.3, **0.047]*** | 55.9 |
| ***TMT-B*** | [0.5, 0.9] | [-3.4, **0.001]*** | [2725.6, **0.0017]*** | [2589.9, **0.013]*** | [3420.3, **0.002**]* | 272.4 |
| ***TMT-BA*** | [2568.9, **0.002]*** | [-2.2, **0.001]*** | [1183.2, 0.2] | [914.5, 0.3] | [2568.9, **0.002**]* | 159.4 |

Table S3.5: **Model 1**, Executive and attentive neuropsychological profile showing significant effects revealed by a linear mixed model of longitudinal changes (pre-surgical and three-months follow-up) in brain tumor patients stratified into two groups (with or without attentional deficit in the immediate post-surgical time point). The model included the following interaction predictors of longitudinal dynamic functional connectivity patterns of the Fronto-Parietal Network: *betweenness centrality* × *attentional deficit* × *time, occurrences* × *attentional deficit* × *time, in degree* × *attentional deficit* × *time, out degree* × *attentional deficit* × *time and resilience* × *attentional deficit* ×*.* For all of them, a positive effect means that longitudinal neuropsychological score increases faster in patients with attentional deficit relative to normal ones during time. The model also included time and attentional deficit presence predictors reported in other tables. Significant ($p < 0.05$) fixed effects are emphasized in bold and an asterisk.

| ***Response*** | ***Predictors*** *[Estimate* β*, p-value]* | | | | | ***Random effects*** |
|---|---|---|---|---|---|---|
| | *Time × Attentional Deficit × Betweenness centrality* | *Time × Attentional Deficit × Occurrences* | *Time × Attentional Deficit × In Degree* | *Time × Attentional Deficit × Out Degree* | *Time × Attentional Deficit × Resilience* | $\sigma^2$ |
| ***Attentional matrices*** | [-1.7, 0.4] | [-0.5, 0.2] | [49.3, 0.9] | [134.4, 0.7] | [562.6, 0.2] | 17.9 |
| ***TMT-A*** | [1.5, 0.7] | [2.9, **0.001]*** | [-1993.2, **0.003]*** | [-2206.3, **0.001]*** | [-2848.1, **0.001]*** | 55.9 |
| ***TMT-B*** | [-15.6, **0.047]*** | [3.7, **0.007]*** | [254.2, 0.9] | [1114.8, 0.4] | [-4518.1, **0.005**]* | 272.4 |
| ***TMT-BA*** | [-16.7, **0.006]*** | [0.8, 0.4] | [1913.7, 0.1] | [3064.5, **0.005]*** | [-1718.8, **0.156**]* | 159.4 |

Table S4: **Model 2**, The presence of attentional deficit showing significant effects revealed by a linear model of dynamic temporal properties of the Fronto-parietal network. The model included the following main predictors of dynamic functional connectivity patterns of the Fronto-Parietal Network: *betweenness centrality; occurrences, in degree, out degree and resilience.* For all of them, a positive effect means neuropsychological score increases regardless of other dynamic properties. Significant ($p < 0.05$) fixed effects are emphasized in bold and an asterisk.

| ***Response*** | **Predictors** *[Estimate β, p-value]* | | | | | |
|---|---|---|---|---|---|---|
| | *Betweeness centrality* | *Occurrences* | *In Degree* | *Out Degree* | *Resilience* | ***$R^2$/ $R^2$ adjusted*** |
| ***Attentional deficit*** | [-0.1, 0.5] | [-0.0, 0.9] | [31.3, **0.0009]*** | [21.7, **0.040]*** | [7.2, 0.5] | 0.2/0.1 |

Table S5: **Model 3**, Pre-surgical neuropsychological scores showing significant effects revealed by a linear model of pre-surgical dynamic temporal properties of the Fronto-parietal network. The model included the following main predictors of dynamic functional connectivity patterns of the Fronto-Parietal Network: *betweenness centrality; occurrences, in degree, out degree and resilience.* For all of them, a positive effect means pre-surgical neuropsychological score increases regardless of other pre-surgical dynamic properties. Significant ($p < 0.05$) fixed effects are emphasized in bold and an asterisk.

| ***Response*** | ***Predictors** [Estimate β, p-value]* | | | | | |
|---|---|---|---|---|---|---|
| | *Betweeness centrality* | *Occurrences* | *In Degree* | *Out Degree* | *Resilience* | ***$R^2$/ $R^2$ adjusted*** |
| ***Attentional matrices*** | [1.6, **0.045]*** | [0.3, **0.035]*** | [-637.6, **0.002]*** | [-517.1, **0.004]*** | [-289.3, 0.09] | 0.2/0.2 |
| ***TMT-A*** | [-3.8, 0.1] | [-1.0, **0.005]*** | [1504.3, **0.003]*]** | [1255.7, **0.005]*** | [1139.9, **0.009]*** | 0.2/0.2 |
| ***TMT-B*** | [-7.3, 0.2] | [-2.5, **0.023]*** | [5322.9, **0.001]*** | [4034.1, **0.003]*** | [2098.2, 0.1] | 0.2/0.2 |
| ***TMT-BA*** | [-3.9, 0.4] | [-1.3, 0.1] | [3716.3, **0.001]*** | [2716.8, **0.005]*** | [820.9, 0.4] | 0.2/0.2 |

Table S6: **Model 4**, Post-surgical neuropsychological scores showing significant effects revealed by a linear model of pre-surgical dynamic temporal properties of the Fronto-parietal network. The model included the following main predictors of dynamic functional connectivity patterns of the Fronto-Parietal Network: *betweenness centrality; occurrences, in degree, out degree and resilience.* For all of them, a positive effect means post-surgical neuropsychological score increases regardless of other pre-surgical dynamic properties. Significant ($p < 0.05$) fixed effects are emphasized in bold and an asterisk.

| ***Response*** | ***Predictors** [Estimate β, p-value]* | | | | | |
|---|---|---|---|---|---|---|
| | *Betweeness centrality* | *Occurrences* | *In Degree* | *Out Degree* | *Resilience* | ***$R^2$/ $R^2$ adjusted*** |
| ***Attentional matrices*** | [0.9, 0.4] | [0.2, 0.3] | [-945.3, **0.001]*** | [-673.9, **0.004]*** | [-12.1, 0.9] | 0.2/0.2 |
| ***TMT-A*** | [-0.4, 0.9] | [0.1, **0.883]*** | [1602.4, **0.038]*** | [1045.4, **0.123]*** | [-637.4, 0.4] | 0.1/0.1 |
| ***TMT-B*** | [-3.9, 0.4] | [-1.3, 0.1] | [5314.1, **0.001]*** | [3716.3, 0.2] | [-2027.5, 0.4] | 0.1/0.7 |
| ***TMT-BA*** | [-1.3, 0.8] | [-0.3, 0.7] | [2848.0, **0.016]*** | [1932.7, 0.1] | [-422.9, 0.7] | 0.2/0.1 |

Table S7: **Model 5**, Three-months follow-up neuropsychological scores showing significant effects revealed by a linear model of pre-surgical dynamic temporal properties of the Fronto-parietal network. The model included the following main predictors of dynamic functional connectivity patterns of the Fronto-Parietal Network: *betweenness centrality; occurrences, in degree, out degree and resilience.* For all of them, a positive effect means follow-up neuropsychological score increases regardless of other pre-surgical dynamic properties. Significant ($p < 0.05$) fixed effects are emphasized in bold and an asterisk.

| ***Response*** | ***Predictors** [Estimate β, p-value]* | | | | | |
|---|---|---|---|---|---|---|
| | *Betweenness centrality* | *Occurrences* | *In Degree* | *Out Degree* | *Resilience* | ***$R^2$/ $R^2$ adjusted*** |
| ***Attentional matrices*** | [1.0, 0.2] | [0.0, 0.9] | [-182.3, 0.4] | [-167.7, 0.4] | [21.8, 0.9] | 0.0/-0.0 |
| ***TMT-A*** | [0.3, 0.8] | [-0.0, 0.9] | [315.3, 0.4] | [192.5, 0.5] | [-96.9, 0.7] | 0.0/-0.0 |
| ***TMT-B*** | [-1.3, 0.8] | [-0.4, 0.7] | [3251.9, **0.020]*** | [2203.1, 0.1] | [-444.4, 0.7] | 0.1/0.1 |
| ***TMT-BA*** | [-1.3, 0.8] | [-0.3, 0.7] | [2848.0, **0.016]*** | [1932.6, 0.1] | [-422.9, 0.7] | 0.1/0.1 |

#### Supplementary Figures

**Figure S1**: A) and B) Significant Latent Component (LC) identified with grouped behavioral Partial Least Square Analysis with all CAPs from the FPN seed explaining differing effects of covariance in patients with different attentive performance disregarding absolute group differences. C) and D) Significant Latent Component (LC) identified with grouped behavioral Partial Least Square Analysis with all CAPs from the FPN seed explaining differing effects of covariance in patients with different attentive performance disregarding absolute group differences.

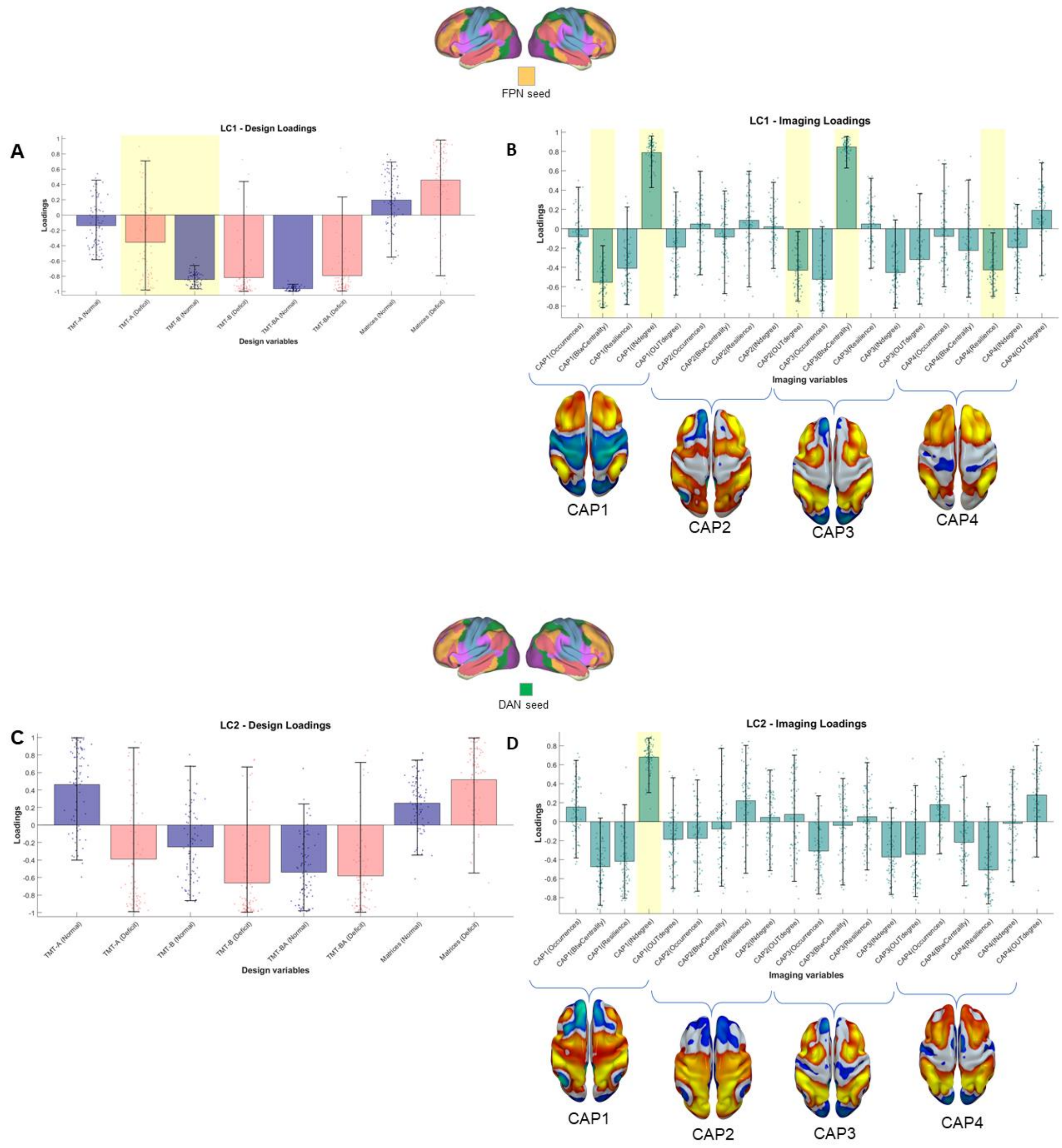

Figure S2: **Prediction of longitudinal attentive/executive scores through longitudinal fronto-parietal temporal properties for Model 1.**

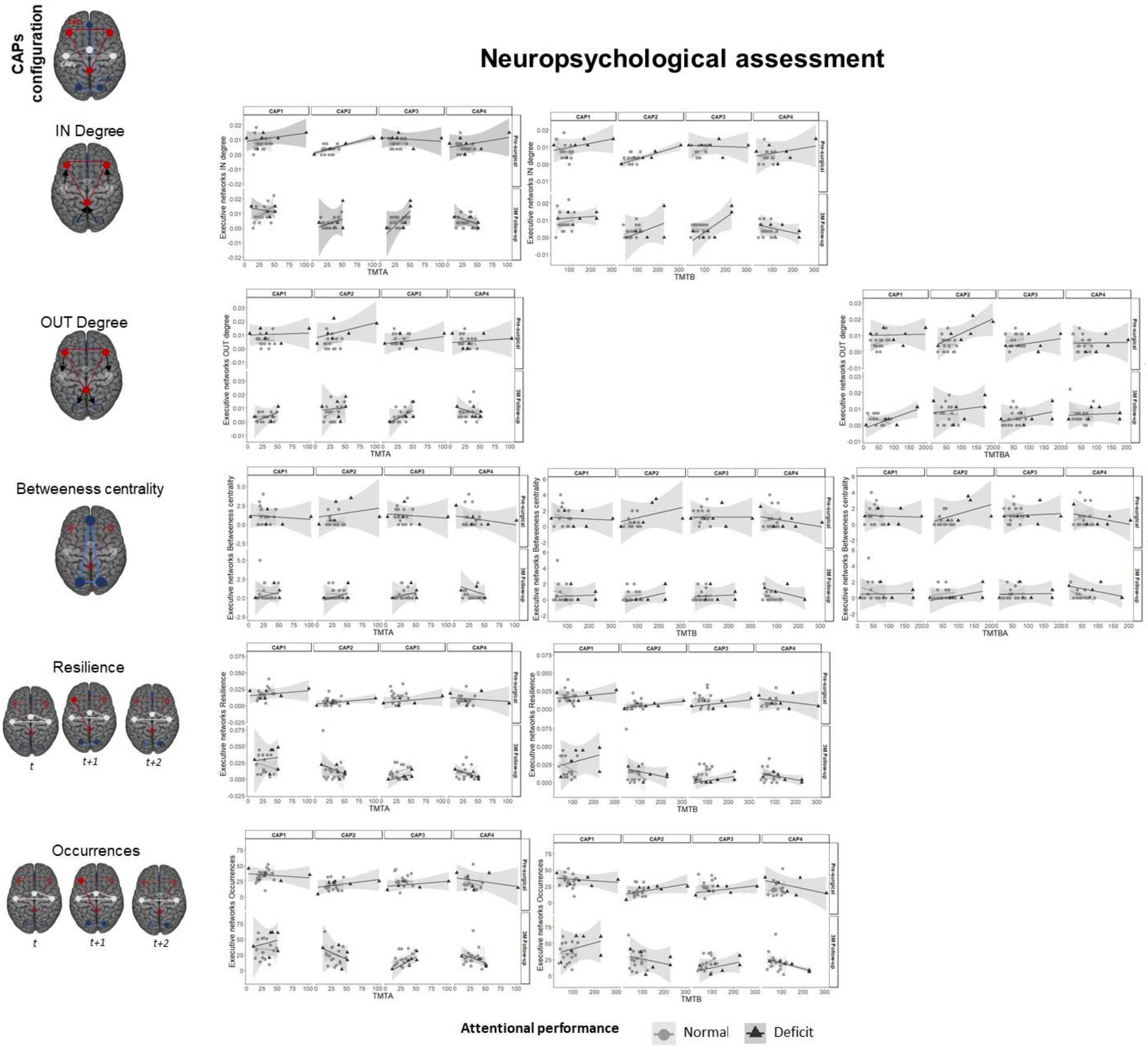
